# Associations of rest-activity rhythm disturbances with stroke risk and post-stroke adverse outcomes

**DOI:** 10.1101/2023.05.14.23289966

**Authors:** Lei Gao, Xi Zheng, Sarah N. Baker, Peng Li, Frank A.J.L. Scheer, Ricardo C Nogueira, Kun Hu

**Author notes:** Address correspondence to: Lei Gao, MBBS; Department of Anesthesia, Massachusetts General Hospital, Boston, MA, 02114; Kun Hu, Ph.D.; Medical Biodynamics Program, Division of Sleep and Circadian Disorders, Brigham and Women’s Hospital, Boston, MA, USA; Twitter: @MBP_Harvard.

## Abstract

**Background:** Almost all biological and disease processes are influenced by circadian clocks and display ∼24-hour rhythms. Disruption of these rhythms may be an important novel risk factor for stroke. We evaluated the association between 24-h rest-activity rhythm measures, stroke risk, and major post-stroke adverse outcomes.

**Methods:** In this cohort study, we examined ∼100,000 participants in the UK Biobank (44-79 years old; ∼57% females) who underwent an actigraphy (6-7 days) and 5-year median follow-up. We derived: (1) most active 10 hours activity counts (*M10*) across the 24-h cycle and the timing of its midpoint (*M10 midpoint*); (2) the least active 5 hours counts (*L5*) and its midpoint timing (*L5 midpoint*); (3) relative amplitude (*RA*) - (M10-L5)/(M10+L5); (4) *interdaily stability* (IS): stability and (5) *intradaily variability* (IV), fragmentation of the rhythm. Cox proportional hazard models were constructed for time to (i) incident stroke (n=1,652); and (ii) post-stroke adverse outcomes (dementia, depression, disability, or death).

**Results:** Suppressed RA (lower M10 and higher L5) was associated with stroke risk after adjusting for demographics; the risk was highest in the lowest quartile [Q1] for RA (HR=1.62; 95% CI:1.36-1.93, *p*<0.001) compared to the top quartile [Q4]. Participants with *later* M10 midpoint timing (14:00-15:26, HR=1.26, CI:1.07-1.49, *p*=0.007) also had a higher risk for stroke than *earlier* (12:17-13:10) participants. A fragmented rhythm (IV) was also associated with a higher risk for stroke (Q4 vs. Q1; HR=1.27; CI:1.06-1.50, *p*=0.008), but differences in the stability of rhythms (IS) were not. Suppressed RA was associated with an increased risk of unfavorable post-stroke outcomes (Q1 vs. Q4; 1.78 [1.29-2.47]; *p*<0.001). All the associations were independent of age, sex, race, obesity, sleep disorders, cardiovascular diseases or risks, and other morbidity burdens.

**Conclusion:** Suppressed 24-h rest-activity rhythm may be a risk factor for stroke and an early indicator of major post-stroke adverse outcomes.

## Introduction

Stroke is a leading cause of mortality and disability,^1^ and it also increases the risks for dementia and depression.^2–4^ How to prevent stroke and improve post-stroke outcomes is still a contemporary challenge.^5^ To adapt to the environmental changes in the day-night cycle, almost all biological/physiological processes from the cellular to system levels (including sleep) display 24-h rhythms.^6,7^ These rhythms are generated and orchestrated by the circadian system, composed of a network of coupled cell-autonomous oscillators or circadian clocks.^8^ Disruption to circadian regulation, commonly seen with shiftwork, is linked to an increased risk for obesity, sleep disorders, hypertension, diabetes, and cardiometabolic disorders^9–14^ — all of which are major risk factors for stroke.^15^ However, few large-scale longitudinal studies have formally examined the association between circadian disruption and risk for incident stroke in the general population.

Advances in wearable technology make it possible to continuously monitor/collect physiological outputs such as motor activity non-invasively for the long term. Indeed, ambulatory motor activity recordings have been widely applied in field studies to estimate circadian rhythms and disturbances. For instance, rest-activity rhythms (RARs) of motor activity based on actigraphy^16^ have been used to demonstrate that daily rhythms are altered with aging and in depressive patients;^17,18^ actigraphy-derived RAR disturbances are associated with amyloid/tau pathology even in cognitively intact humans;^19,20^ and predict cognitive decline, Alzheimer’s disease (AD) and related dementias, depression, disability, and mortality.^19,21–26^ In addition, studies have revealed altered circadian and sleep-wake patterns (via RARs) in patients with stroke.^27–29^ Given that increased risk for dementia, depression, disability, and mortality are well-established long-term adverse outcomes in post-stroke survivors,^30,31^ understanding the relationship between altered RARs and such post-stroke outcomes is vital for prevention and long-term management.

To understand the relationship between circadian disturbances, incident stroke risk, and post-stroke outcomes, we examined RARs of motor activity (actigraphy) collected from over 100,000 middle-older aged adults in the UK Biobank between 2013 and 2015. After the actigraphy assessment at baseline, participants’ health was followed via electronic health records for up to 7.5 years. We tested two hypotheses: 1) participants with more RARs disturbances at baseline will have a higher risk for incident stroke, and 2) these RARs disturbances predict post-stroke adverse outcomes (i.e., new-onset depression, disability, dementia, or death).

## Methods

### Study Population and Data Source

This study involved the analysis of data collected from the participants in the UK Biobank (UKB) — a longitudinal population-based cohort (age range: 43-79 years old; 54% female). UKB participants completed extensive questionnaires on demographics, lifestyle choices, and medical conditions at initial enrollment. They agreed to provide access to their healthcare records collected in the UK Biobank’s Hospital Inpatient Data, an electronic health record for each participant from the UK’s centralized National Health Service (NHS).^32^ This study focused on 103,711 UKB participants who agreed to have actigraphy assessment between 2013 and 2015 (2.8 to 9.7 years after enrollment) — baseline for the current study. Follow-up for participants was until September 2021, up to 7.5 (median 5) years from baseline.

To address the primary hypothesis (H 1) that circadian disturbances at baseline were associated with an increased risk of stroke, we included only those participants who had no stroke (any type) or transient ischemic attack before baseline (Fig. 1). To test the second hypothesis (H2) that circadian disturbances predicted post-stroke adverse outcomes, dementia, depression, disability, or death (four competing outcome measures), we included only participants with stroke by the time of actigraphy assessment.

**Figure 1.**
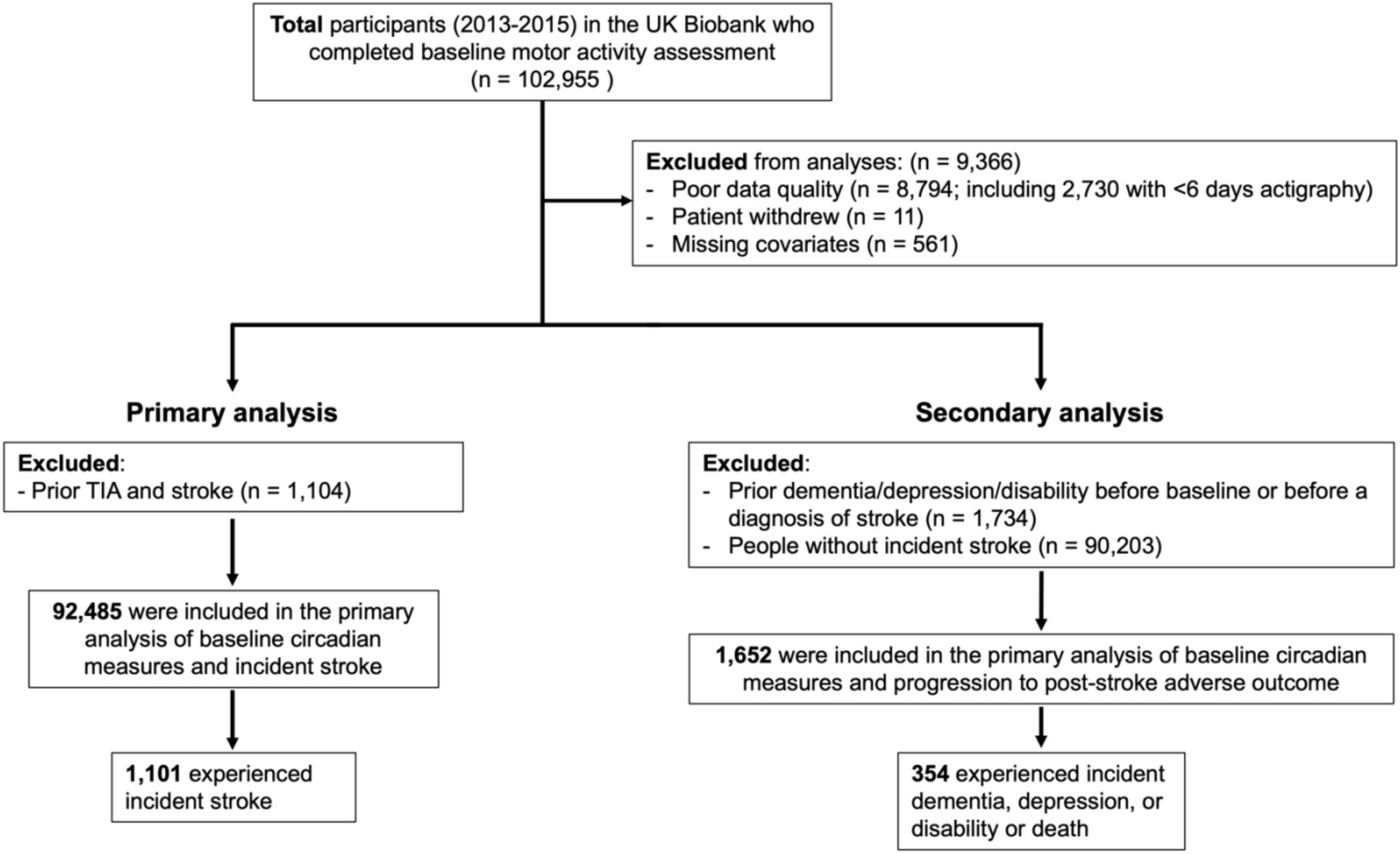
Flowchart of participants in the study. The primary analysis included 1,101 incident strokes. 1,632 stroke cases without dementia, depression, or disability were included in the secondary analysis. *TIA* transient ischemic attack.

### Standard Protocol Approvals, Registrations, and Patient Consents

The UK Biobank received National Research Ethics Approval, and participants gave written informed consent. This study was conducted under the terms of UK Biobank access number 33883 and Mass General Brigham IRB approval (#2020P002097).

### Assessment of Rest-Activity Rhythms (RARs)

A triaxial accelerometer device (Axivity AX3) was worn continuously for up to 7 days. Data collection and quality checks were performed based on prior work with actigraphy in older adults^20,22^ and established criteria from the UK Biobank.^33^ Accelerometer data were sampled at ∼100 Hz and used to derive activity counts in each 15-sec epoch.

To quantify circadian rest-activity rhythms, we analyzed each recording of activity counts using non-parametric analyses and derived the following RAR measures:^34^ (1) *M10* activity counts of the most active 10 hours and the timing of its midpoint (*M10 midpoint*) across the 24-h cycles that represent physical activity level and the timing of the active phase, respectively; (2) *L5* activity counts of the least active 5 hours and the timing of its midpoint (*L5 midpoint*) that represent the level of movement or wake-up and the timing of the “nocturnal” (most likely sleep) period, respectively; (3) the relative amplitude (RA) based on activity counts for M10 and L5, (M10-L5)/(M10+L5) that estimates the robustness of the 24-h rhythm; (4) *interdaily stability* (IS) for assessment of the stability of the rhythm across different days (i.e., a larger IS represents a more stable rhythm); and (5) *intradaily variability* (IV) for assessment of fragmentation of the rhythm (i.e., a smaller IV represents less fragmentation of the rhythm).

We excluded 8,794 records (8.5%) due to flagged cases with data problems, poor calibration, and large gaps/likely off-wrist periods. For the analysis of RAR, we also excluded those recordings with <6 days (n = 2,730; see Fig. 1); for those recordings longer than 6 days, we used the first 6 available days of data to ensure the same data length that might affect certain RAR measures such as IS and IV. All the RAR analyses were performed using software implemented in MATLAB (Ver. R2020a, The MathWorks Inc., Natick, MA, USA).

### Assessment of stroke and post-stroke adverse outcomes

The UK Biobank has released hospitalization records linked to study participants during the follow-up period within the UK’s NHS. Incident stroke diagnosis was derived as the first date of occurrence of the ICD-10 codes (ICD-10: H341, I63, I693, G46, I60, I61, I62, I690, I691, I692, I64, I694), included in hospital admissions health records during follow-up, in keeping with similar studies using these data.^35^ In addition, we considered ischemic stroke and hemorrhagic stroke separately. We obtained incident depression/affective disorder (ICD-10: F32, F33, F34, F38, F39), disability diagnosis (ICD-10: R54, Z736), and dementia diagnoses using the UK Biobank’s algorithmically defined “date of all-cause dementia” (field 42018). The UK Biobank released the date of death via death certificates within the UK National Health Service (NHS). Time-to-event (first occurrence of dementia, depression, disability, or death) was derived using the first date of diagnosis and the date of actigraphy assessment.

### Assessment of Covariates

The UK Biobank made participants’ medical histories available through a combination of self-reports during nurse-led interviews at initial enrollment or from medical records at the time of actigraphy. Covariates were grouped into four categories. (1) Demographics include age, sex, ethnicity, education, and deprivation): age at actigraphy was calculated in years based on the date of birth, sex (male/female), and ethnicity [European/non-European, given that the majority was of British or “white” European descent (96.6%)] were self-reported; Education was college-level (yes/no); and Townsend deprivation index (TDI) was a score based on national geographic census data immediately before participant enrollment. (2) Sleep disorders were determined based on ICD-10 codes (F51 and G47). (3) Total nighttime duration [TST]) was derived from actigraphy consistent with our prior studies^22,36^ and was divided into three groups (<7h, 7-8h, and >8h) (4) Comorbidities include body mass index (BMI)>30, presence of cardiovascular disease or risk factors (CVD), and morbidity burden. BMI at initial enrollment was calculated as weight [kg] divided by height squared [m^2^]. CVD/risk was based on the presence of hypertension, high cholesterol, being a current smoker, diabetes, ischemic heart disease, and peripheral vascular disease. We also used a previously described morbidity burden based on the summed presence of any cancers, respiratory, neurological, gastrointestinal, renal, hematological, endocrine, musculoskeletal, connective tissue, and infectious diseases/disorders at the time of actigraphy.^23,37–39^

### Statistical analysis

Descriptive characteristics are presented as means with SDs for quantitative variables (normally distributed) and medians with interquartile range (non-normally distributed). Categorical variables are presented as percentages. Cox proportional hazard models were performed to address the proposed two hypotheses, and results were reported as hazard ratios (HRs) and corresponding 95% confidence intervals (CIs). Participants were divided into four quartiles (Q1-Q4) for each RAR measure. For reference levels, we used the highest quartile (Q4) for relative amplitude (i.e., most robust rhythms) and IS (i.e., most stable rhythms) and the lowest quartile (Q1) for IV (i.e., least fragmented rhythms) based on prior findings.^23,40^ Given the 24h clock, we categorized M10 and L5 midpoints starting from a median time and defined a “middle” quartile (with its associated boundary clock times). The quartiles before and after were defined as “earlier” and “later.” The final quartile of participants beyond these quartile boundary times was defined as “extremes.” M10 midpoint periods are earlier: 12:17-13:10, middle: 13:11-13:59, later: 14:00-15:26, and extremes: <12:17, >15:26. L5 midpoint periods are earlier: 2:04-2:56, middle: 2:57-3:39, later: 3:40-4:38, extremes <2:04, >4:38 (see eFig. 1). We hypothesized that “earlier” M10 midpoint, (corresponding to earlier activities during the day i.e., morning/early afternoon) and “earlier” L5 midpoint, (corresponding to an earlier preference for going to bed), represented the lowest risk groups and were used as reference levels.

(1) Primary analysis was focused on the associations between RAR measures and stroke incidence in those participants without stroke at baseline (hypothesis 1). 92,485 participants (baseline age: mean±SD 62.4±7.8 years old, range 43.5-78.7 years; female: 56.6%) with valid actigraphy recordings and covariates were used in the analysis, representing over half a million person days of actigraphy. The core models included M10, L5, M10 midpoint, L5 midpoint, RA, IS, and IV (quartiles) as predictors alongside demographics. Adjusted models were subsequently used to further control for the other categories of covariates. The same core and fully adjusted models were repeated for all strokes and ischemic and hemorrhagic strokes separately.

(2) Secondary analysis was focused on the associations between RAR measures and adverse post-stroke outcomes (depression, disability, dementia, and death) (hypothesis 2). After excluding stroke patients with pre-existing dementia, depression, or disability, a total of 1,652 stroke patients (baseline age: mean±SD 66.7±6.8 years old, range 46.6-77.8 years; female: 38.7%) were included in the analysis. Each RAR measure was included separately as a predictor alongside demographics and time lag from stroke diagnosis to actigraphy assessment (core models). Fully adjusted models included all other categories of covariates (obesity, sleep disorders, cardiovascular diseases or risks, and other morbidity burdens). Missing covariate data was <1% (Fig. 1) and excluded from the models. All statistical analyses were performed using JMP Pro (Ver. 16, SAS Institute, Cary, NC, USA). *p*-value < 0.05 was used for statistical significance.

### Data availability

Data are available from the UK Biobank after applying (https://www.ukbiobank.ac.uk/enable-your-research/apply-for-access). All scripts for conducting the analysis are available upon reasonable request.

## Result

### Participant characteristics

Baseline demographics, daily rhythmicity characteristics, sleep, physical activity, and participants’ comorbidities for two hypotheses are shown in Table 1. In those participants without stroke at baseline (for Hypothesis 1), 1101 participants (out of 92,485) had their first stroke event during a median 3.9-year follow-up. Within those stroke events after baseline, 765 were ischemic strokes, and 290 were hemorrhagic strokes (46 other stroke events were unspecified). Of stroke patients, 354 (out of 1,652) were newly diagnosed with depression, disability, or dementia or died during a median 4.5-year follow-up (range 1.2 months to 8.1 years; SD 1.9 years) after the baseline.

**Table1.**
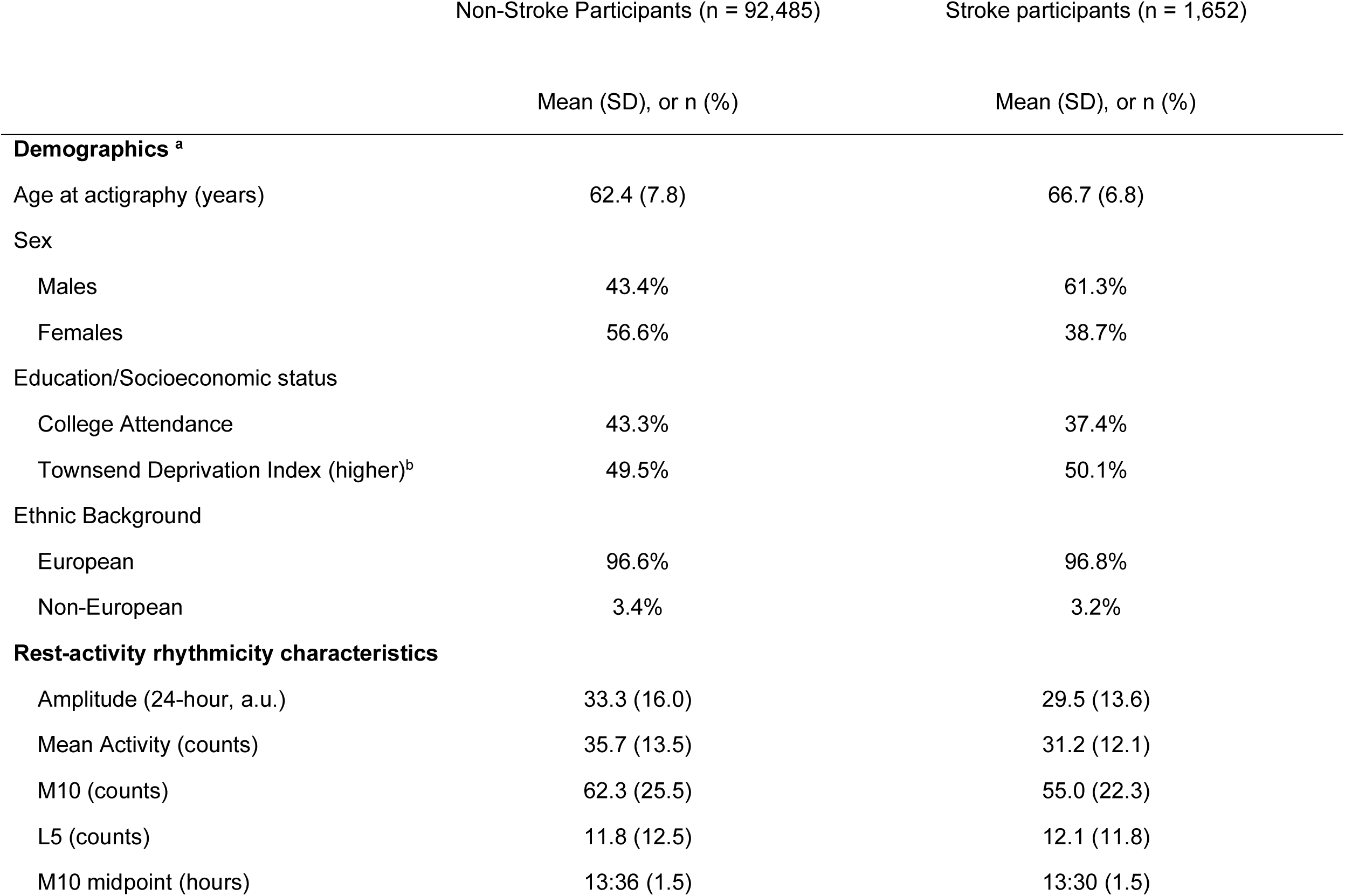
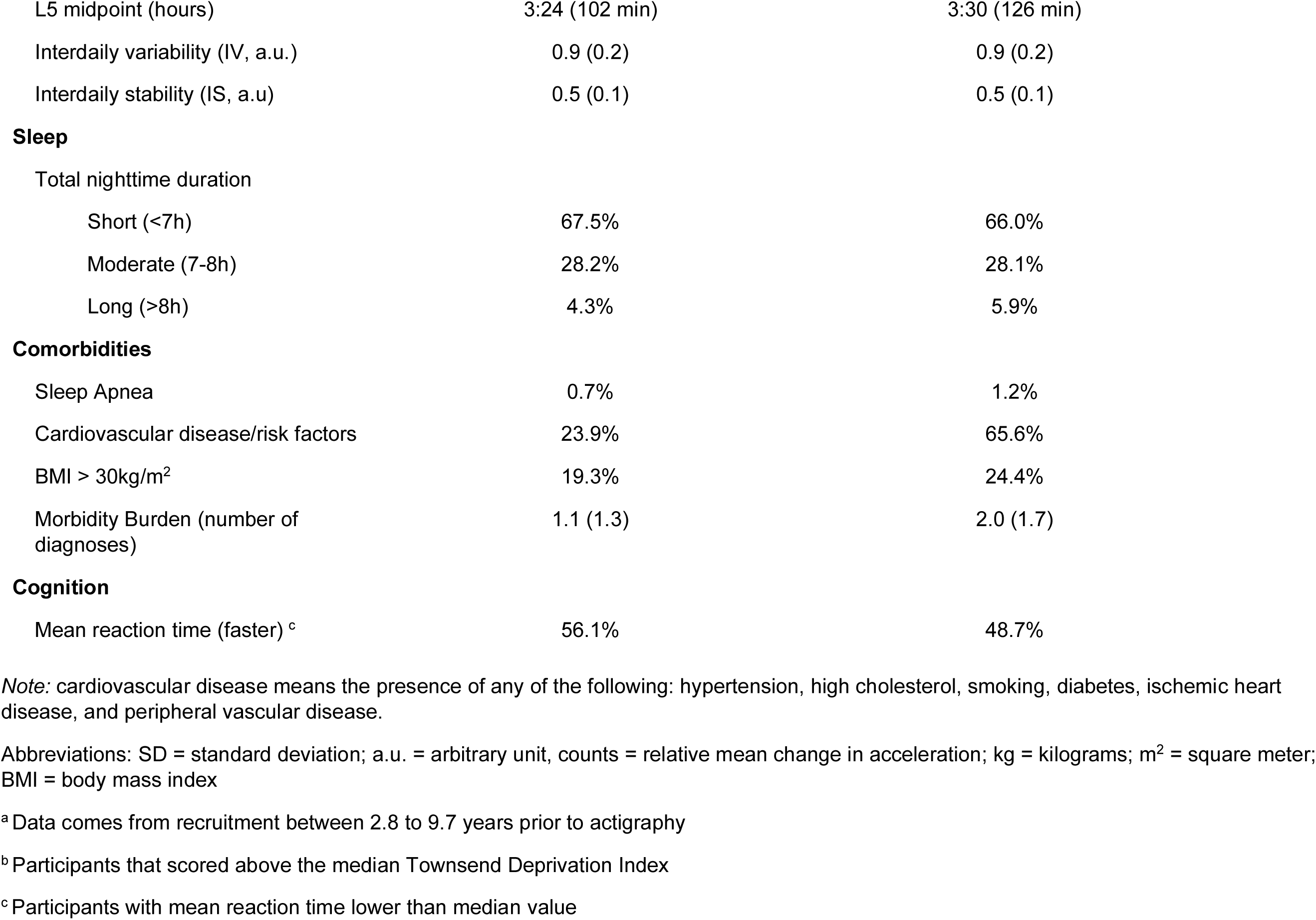
Demographics, lifestyle, and clinical comorbidities.

|  | Non-Stroke Participants (n = 92,485) | Stroke participants (n = 1,652) |
| --- | --- | --- |
|  | Mean (SD), or n (%) | Mean (SD), or n (%) |
| **Demographics <sup>a</sup>** |  |  |
| Age at actigraphy (years) | 62.4 (7.8) | 66.7 (6.8) |
| Sex |  |  |
| Males | 43.4% | 61.3% |
| Females | 56.6% | 38.7% |
| Education/Socioeconomic status |  |  |
| College Attendance | 43.3% | 37.4% |
| Townsend Deprivation Index (higher)<sup>b</sup> | 49.5% | 50.1% |
| Ethnic Background |  |  |
| European | 96.6% | 96.8% |
| Non-European | 3.4% | 3.2% |
| **Rest-activity rhythmicity characteristics** |  |  |
| Amplitude (24-hour, a.u.) | 33.3 (16.0) | 29.5 (13.6) |
| Mean Activity (counts) | 35.7 (13.5) | 31.2 (12.1) |
| M10 (counts) | 62.3 (25.5) | 55.0 (22.3) |
| L5 (counts) | 11.8 (12.5) | 12.1 (11.8) |
| M10 midpoint (hours) | 13:36 (1.5) | 13:30 (1.5) |
| L5 midpoint (hours) | 3:24 (102 min) | 3:30 (126 min) |
| Interdaily variability (IV, a.u.) | 0.9 (0.2) | 0.9 (0.2) |
| Interdaily stability (IS, a.u) | 0.5 (0.1) | 0.5 (0.1) |
| **Sleep** |  |  |
| Total nighttime duration |  |  |
| Short (<7h) | 67.5% | 66.0% |
| Moderate (7-8h) | 28.2% | 28.1% |
| Long (>8h) | 4.3% | 5.9% |
| **Comorbidities** |  |  |
| Sleep Apnea | 0.7% | 1.2% |
| Cardiovascular disease/risk factors | 23.9% | 65.6% |
| BMI > 30kg/m<sup>2</sup> | 19.3% | 24.4% |
| Morbidity Burden (number of diagnoses) | 1.1 (1.3) | 2.0 (1.7) |
| **Cognition** |  |  |
| Mean reaction time (faster) <sup>c</sup> | 56.1% | 48.7% |

*Note:* cardiovascular disease means the presence of any of the following: hypertension, high cholesterol, smoking, diabetes, ischemic heart disease, and peripheral vascular disease.
Abbreviations: SD = standard deviation; a.u. = arbitrary unit, counts = relative mean change in acceleration; kg = kilograms; m<sup>2</sup> = square meter; BMI = body mass index
<sup>a</sup> Data comes from recruitment between 2.8 to 9.7 years prior to actigraphy
<sup>b</sup> Participants that scored above the median Townsend Deprivation Index
<sup>c</sup> Participants with mean reaction time lower than median value

### Rest-activity rhythms and incident stroke risk

Lower RA was associated with a higher risk for stroke after adjusting for demographic covariates (*p*<0.001; Table 2; Fig. 2A). Compared to those in the top quartile [Q4], the risk for developing stroke was higher in the third quartile [Q3] (hazard ratio [HR]: 1.25, 95% confidence interval [CI]: 1.04-1.51, *p*=0.018), second quartile [Q2] (HR=1.40, 95% CI:1.17-1.68, *p*<0.001), and the lowest quartile [Q1] (HR=1.62; 95% CI: 1.36-1.93, *p*<0.001). Consistent with the findings for RA, lower M10 (Q1 vs. Q4 core model: *p*<0.001) and higher L5 (Q4 vs. Q1 core model: *p*<0.01) were also associated with increased risk for stroke after adjusting for demographic covariates; L5 results were attenuated in the fully adjusted model (Table 2; Fig. 2B-C).

**Figure 2.**
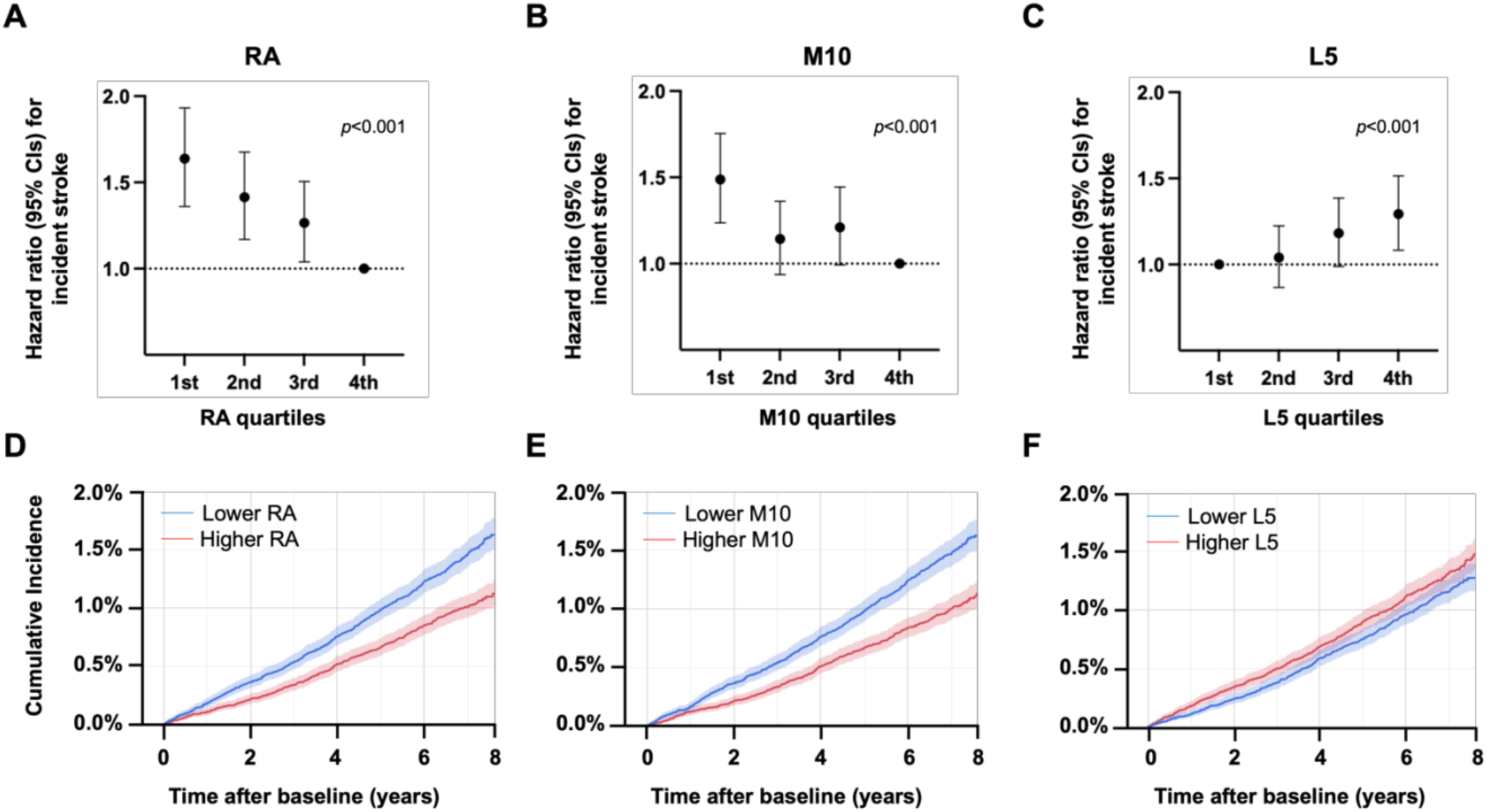
Rest-activity rhythm and risk for stroke. Hazard ratio for stroke risk for (**A**) relative amplitude (***RA***) quartiles with the 4th quartile (the lowest amplitude) as the reference, (**B**) M10 quartiles with the 4th quartile as the reference, and (**C**) L5 quartiles with the 1st quartile (the highest level) as the reference. The cumulative incidence for stroke since baseline (actigraphy assessment) for (**D**) patients with lower RA (1st and 2nd quartiles) and higher RA (3rd and 4th quartiles), (**E**) patients with lower M10 (1st and 2nd quartiles) and higher M10 (3rd and 4th quartiles), and (**F**) patients with lower L5 (1st and 2nd quartiles) and higher L5 (3rd and 4th quartiles). Results were obtained for the first occurrence of stroke adjusted for age, sex, education, ethnicity, and Townsend Deprivation Index. M10: activity level during the most active 10 hours, L5: activity level during the least active 5 hours, ***CI***: confidence interval.

**Table 2.**
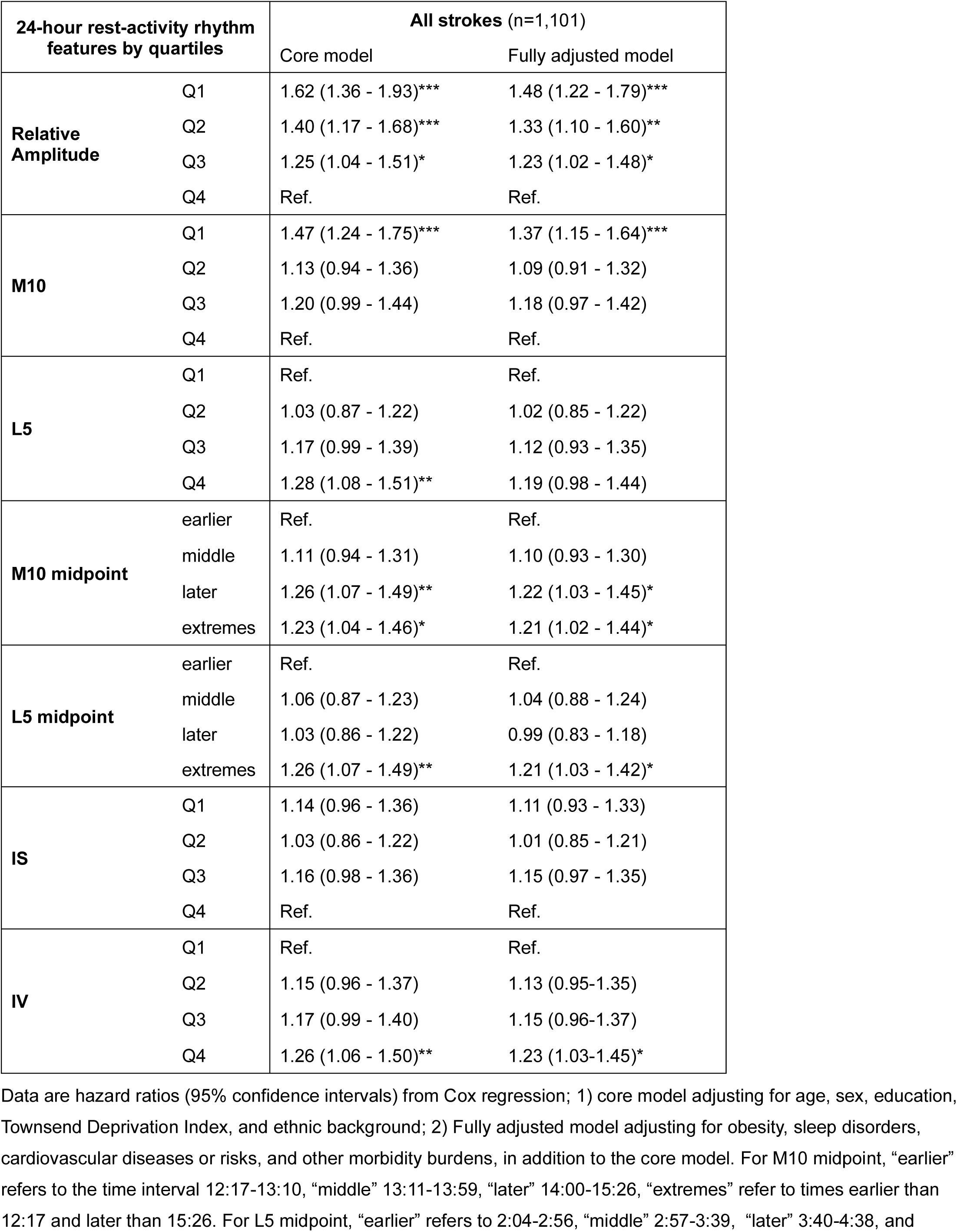
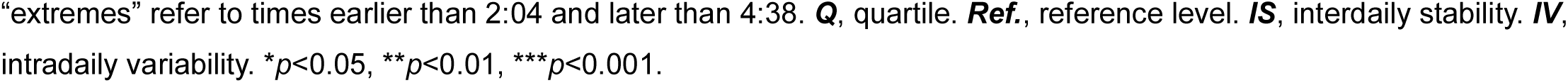
Association between 24-hour rest-activity rhythm (RAR) features and risk for incident stroke.

| 24-hour rest-activity rhythm features by quartiles |  | All strokes (n=1,101) |  |
| --- | --- | --- | --- |
|  |  | Core model | Fully adjusted model |
| **Relative Amplitude** | Q1 | 1.62 (1.36 - 1.93)*** | 1.48 (1.22 - 1.79)*** |
|  | Q2 | 1.40 (1.17 - 1.68)*** | 1.33 (1.10 - 1.60)** |
|  | Q3 | 1.25 (1.04 - 1.51)* | 1.23 (1.02 - 1.48)* |
|  | Q4 | Ref. | Ref. |
| **M10** | Q1 | 1.47 (1.24 - 1.75)*** | 1.37 (1.15 - 1.64)*** |
|  | Q2 | 1.13 (0.94 - 1.36) | 1.09 (0.91 - 1.32) |
|  | Q3 | 1.20 (0.99 - 1.44) | 1.18 (0.97 - 1.42) |
|  | Q4 | Ref. | Ref. |
| **L5** | Q1 | Ref. | Ref. |
|  | Q2 | 1.03 (0.87 - 1.22) | 1.02 (0.85 - 1.22) |
|  | Q3 | 1.17 (0.99 - 1.39) | 1.12 (0.93 - 1.35) |
|  | Q4 | 1.28 (1.08 - 1.51)** | 1.19 (0.98 - 1.44) |
| **M10 midpoint** | earlier | Ref. | Ref. |
|  | middle | 1.11 (0.94 - 1.31) | 1.10 (0.93 - 1.30) |
|  | later | 1.26 (1.07 - 1.49)** | 1.22 (1.03 - 1.45)* |
|  | extremes | 1.23 (1.04 - 1.46)* | 1.21 (1.02 - 1.44)* |
| **L5 midpoint** | earlier | Ref. | Ref. |
|  | middle | 1.06 (0.87 - 1.23) | 1.04 (0.88 - 1.24) |
|  | later | 1.03 (0.86 - 1.22) | 0.99 (0.83 - 1.18) |
|  | extremes | 1.26 (1.07 - 1.49)** | 1.21 (1.03 - 1.42)* |
| **IS** | Q1 | 1.14 (0.96 - 1.36) | 1.11 (0.93 - 1.33) |
|  | Q2 | 1.03 (0.86 - 1.22) | 1.01 (0.85 - 1.21) |
|  | Q3 | 1.16 (0.98 - 1.36) | 1.15 (0.97 - 1.35) |
|  | Q4 | Ref. | Ref. |
| **IV** | Q1 | Ref. | Ref. |
|  | Q2 | 1.15 (0.96 - 1.37) | 1.13 (0.95-1.35) |
|  | Q3 | 1.17 (0.99 - 1.40) | 1.15 (0.96-1.37) |
|  | Q4 | 1.26 (1.06 - 1.50)** | 1.23 (1.03-1.45)* |

Data are hazard ratios (95% confidence intervals) from Cox regression; 1) core model adjusting for age, sex, education, Townsend Deprivation Index, and ethnic background; 2) Fully adjusted model adjusting for obesity, sleep disorders, cardiovascular diseases or risks, and other morbidity burdens, in addition to the core model. For M10 midpoint, “earlier” refers to the time interval 12:17-13:10, “middle” 13:11-13:59, “later” 14:00-15:26, “extremes” refer to times earlier than 12:17 and later than 15:26. For L5 midpoint, “earlier” refers to 2:04-2:56, “middle” 2:57-3:39, “later” 3:40-4:38, and
“extremes” refer to times earlier than 2:04 and later than 4:38. **Q**, quartile. **Ref.**, reference level. **IS**, interdaily stability. **IV**, intradaily variability. \* $p < 0.05$ , \*\* $p < 0.01$ , \*\*\* $p < 0.001$ .

M10 midpoint occurring “later” (HR=1.26, 95% CI: 1.07-1.49, *p*=0.007) or during “extreme” times (HR=1.23, CI: 1.04-1.46, *p*=0.02) had a higher risk for stroke compared to early participants (Fig. 3A). Only “extremes” in the L5 midpoint timing (HR=1.26, 95% CI: 1.07-1.49, *p*=0.005) had a higher risk for stroke compared to early participants (Fig. 3B). A more fragmented rhythm (IV Q4 vs. Q1; HR=1.26, 95% CI: 1.06-1.50, *p*=0.008) was associated with a higher risk for stroke, but the stability of rhythms (IS) was not (Table 2). These associations were consistent for ischemic stroke, but no significant associations were observed between RAR measures and hemorrhagic stroke (eTable 1).

**Figure 3.**
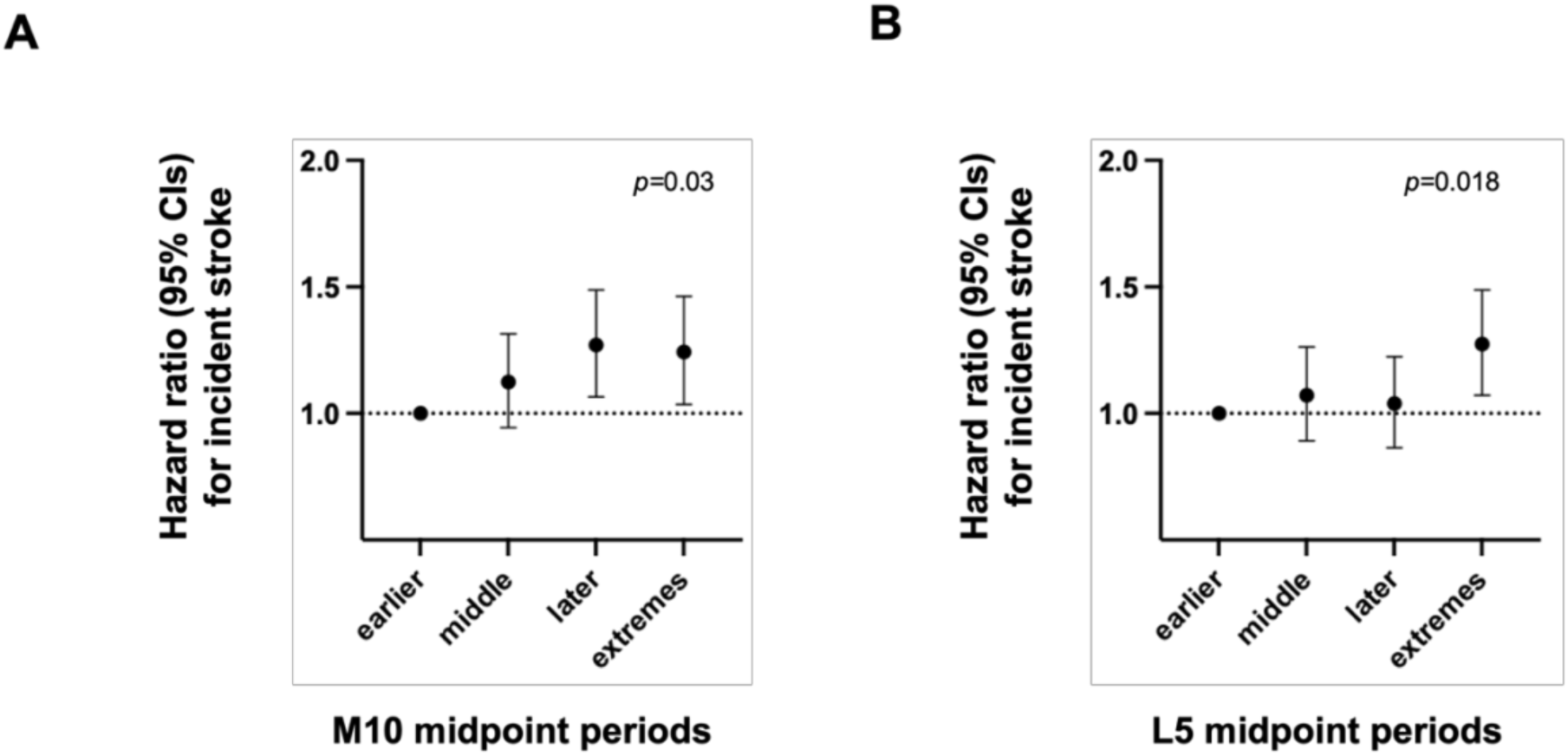
Timing of rest-activity rhythm and incident stroke risk. Hazard ratios for stroke risk for (A) M10 midpoint quartiles with the “earlier” quartile (12:17-13:10) as the reference, (B) L5 midpoint quartiles with the “earlier” quartile (2:04-2:56) as the reference. ***CI:*** confidence interval.

### Rest-activity rhythms and post-stroke adverse outcomes

In participants with any stroke, lower RA [Q1 vs. Q4] was associated with higher risk (HR: 1.78; 95% CI: 1.29-2.47, *p*<0.001) for a post-stroke adverse outcome after adjusting for demographic covariates and the time lag between stroke and actigraphy assessment (Table 3 & eFig. 2). Lower M10 [Q1 vs. Q4] was also associated with a higher risk (HR: 1.57; 95% CI: 1.13-2.18, *p*=0.008). However, no other RAR measures were significantly associated with the risk for post-stroke adverse outcomes.

**Table 3.**
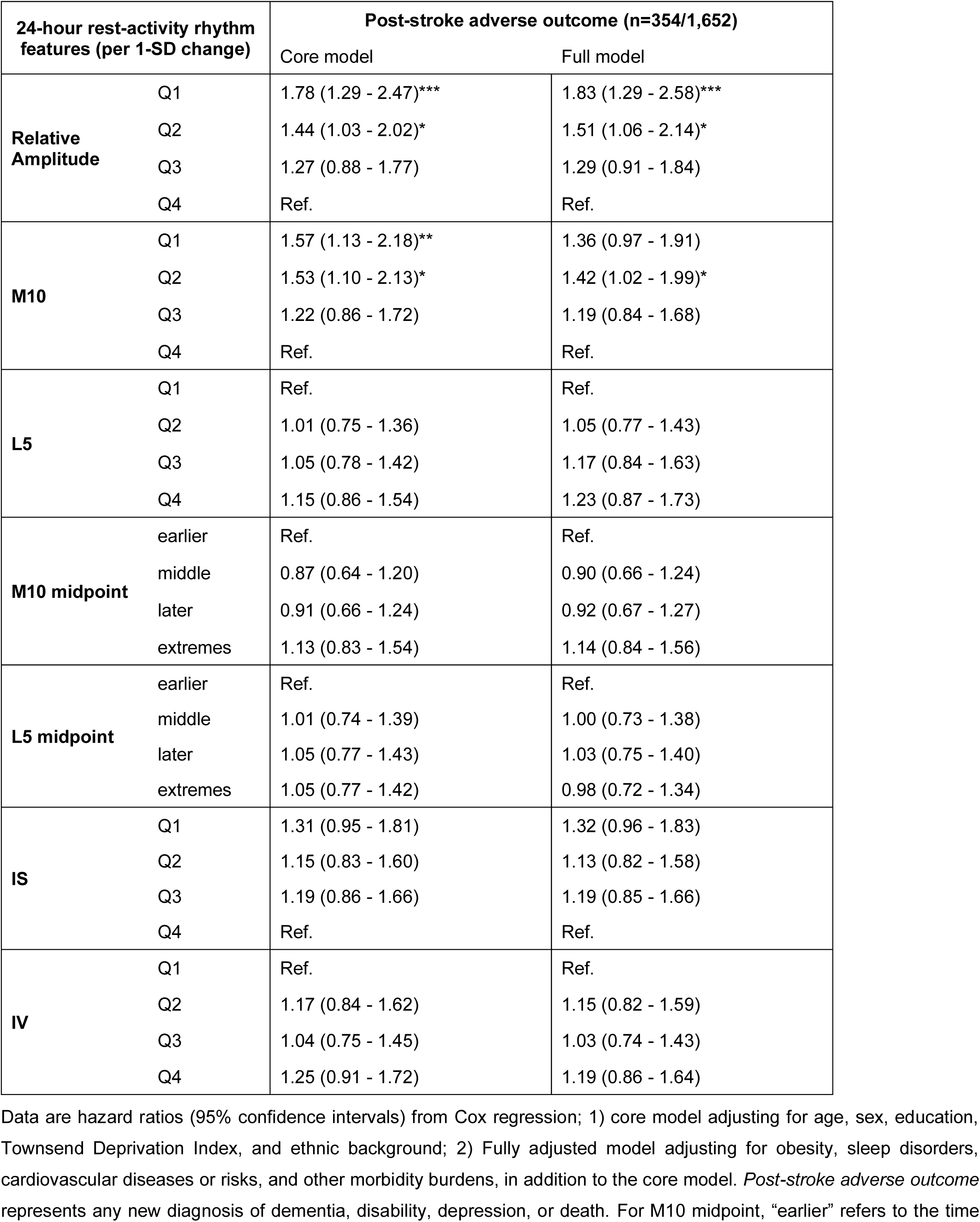
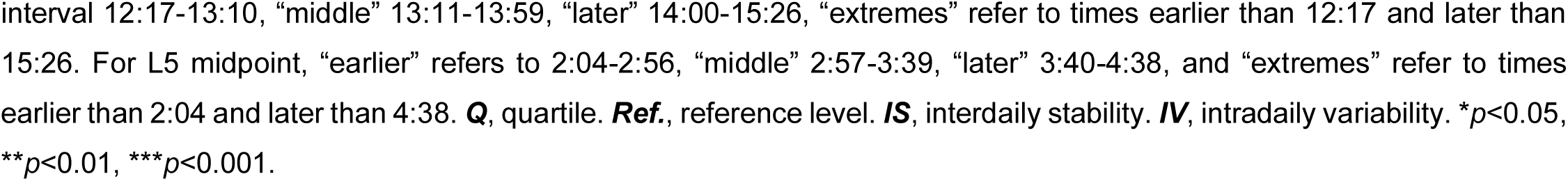
Risk for post-stroke depression, dementia, disability, or death related to baseline 24-hour rest-activity rhythm (RAR) features.

| 24-hour rest-activity rhythm features (per 1-SD change) |  | Post-stroke adverse outcome (n=354/1,652) |  |
| --- | --- | --- | --- |
|  |  | Core model | Full model |
| **Relative Amplitude** | Q1 | 1.78 (1.29 - 2.47)*** | 1.83 (1.29 - 2.58)*** |
|  | Q2 | 1.44 (1.03 - 2.02)* | 1.51 (1.06 - 2.14)* |
|  | Q3 | 1.27 (0.88 - 1.77) | 1.29 (0.91 - 1.84) |
|  | Q4 | Ref. | Ref. |
| **M10** | Q1 | 1.57 (1.13 - 2.18)** | 1.36 (0.97 - 1.91) |
|  | Q2 | 1.53 (1.10 - 2.13)* | 1.42 (1.02 - 1.99)* |
|  | Q3 | 1.22 (0.86 - 1.72) | 1.19 (0.84 - 1.68) |
|  | Q4 | Ref. | Ref. |
| **L5** | Q1 | Ref. | Ref. |
|  | Q2 | 1.01 (0.75 - 1.36) | 1.05 (0.77 - 1.43) |
|  | Q3 | 1.05 (0.78 - 1.42) | 1.17 (0.84 - 1.63) |
|  | Q4 | 1.15 (0.86 - 1.54) | 1.23 (0.87 - 1.73) |
| **M10 midpoint** | earlier | Ref. | Ref. |
|  | middle | 0.87 (0.64 - 1.20) | 0.90 (0.66 - 1.24) |
|  | later | 0.91 (0.66 - 1.24) | 0.92 (0.67 - 1.27) |
|  | extremes | 1.13 (0.83 - 1.54) | 1.14 (0.84 - 1.56) |
| **L5 midpoint** | earlier | Ref. | Ref. |
|  | middle | 1.01 (0.74 - 1.39) | 1.00 (0.73 - 1.38) |
|  | later | 1.05 (0.77 - 1.43) | 1.03 (0.75 - 1.40) |
|  | extremes | 1.05 (0.77 - 1.42) | 0.98 (0.72 - 1.34) |
| **IS** | Q1 | 1.31 (0.95 - 1.81) | 1.32 (0.96 - 1.83) |
|  | Q2 | 1.15 (0.83 - 1.60) | 1.13 (0.82 - 1.58) |
|  | Q3 | 1.19 (0.86 - 1.66) | 1.19 (0.85 - 1.66) |
|  | Q4 | Ref. | Ref. |
| **IV** | Q1 | Ref. | Ref. |
|  | Q2 | 1.17 (0.84 - 1.62) | 1.15 (0.82 - 1.59) |
|  | Q3 | 1.04 (0.75 - 1.45) | 1.03 (0.74 - 1.43) |
|  | Q4 | 1.25 (0.91 - 1.72) | 1.19 (0.86 - 1.64) |

Data are hazard ratios (95% confidence intervals) from Cox regression; 1) core model adjusting for age, sex, education, Townsend Deprivation Index, and ethnic background; 2) Fully adjusted model adjusting for obesity, sleep disorders, cardiovascular diseases or risks, and other morbidity burdens, in addition to the core model. *Post-stroke adverse outcome* represents any new diagnosis of dementia, disability, depression, or death. For M10 midpoint, “earlier” refers to the time
interval 12:17-13:10, “middle” 13:11-13:59, “later” 14:00-15:26, “extremes” refer to times earlier than 12:17 and later than 15:26. For L5 midpoint, “earlier” refers to 2:04-2:56, “middle” 2:57-3:39, “later” 3:40-4:38, and “extremes” refer to times earlier than 2:04 and later than 4:38. **Q**, quartile. **Ref.**, reference level. **IS**, interdaily stability. **IV**, intradaily variability. \* $p<0.05$ , \*\* $p<0.01$ , \*\*\* $p<0.001$ .

## Discussion

### Summary

This is the first large-scale prospective study examining the direct link between rest-activity rhythms and risk for stroke. The key finding is that suppressed 24-h rest-activity rhythms predicted a higher risk of stroke many years before the incident, independent of previously identified risk factors for stroke, including age, sex, race, obesity, sleep disorders, cardiovascular diseases/risks (hypertension, high cholesterol, smoking, diabetes, ischemic heart disease, and peripheral vascular disease) and overall morbidity burden. In addition, a more suppressed 24-h rest-activity rhythm predicted worse long-term outcomes post-stroke. This study presents compelling evidence that daily rhythm disturbances have implications for stroke prevention and recovery.

### Circadian disturbance as a modifiable risk factor for stroke

Circadian disturbances occur when external stimuli such as light,^41^ physical activity,^42,43^ food,^44^ and temperature^45^ provide conflicting circadian time cues^46^, as occurred during shift work,^47^ leading to misaligned, suppressed or abolished daily/circadian rhythms of behavior and physiology. It is known that chronic shift workers experience a higher stroke incidence and risk for other vascular events.^48^ The observed association of suppressed 24-h activity rhythms (lower RA) with increased risk for stroke provides direct evidence for 24-h rhythm disturbances as a modifiable risk factor. In addition to work schedule, many other lifestyle factors such as daytime inactivity, extended nighttime light exposure, social jet lag, and irregular meal timing can temporarily disrupt ‘normal’ daily behavioral rhythms (i.e., acute effect), and chronic exposure to these external factors can disrupt the circadian/sleep control system.^49,50^ These results support the notion that 24-h rhythm disturbances impact stroke risk in a large proportion of the population.

The observed effects of RAR measures (RA, M10) on stroke risk may (partially) overlap with the previously reported effect of exercise on stroke risk^51^ while providing complementary information. RA increases with higher M10 (physical activity level during the most activity 10 hours across 24 h) and lower L5 (nocturnal activity). M10 depends on the intensity, duration, and timing of behavioral activity. However, the benefit of exercise/physical activity during the active period (M10) may be coupled with improved sleep or rest during the inactive period (L5). Similarly, this pattern can be seen in less fragmented sleep-wake cycles (lower IV). Only individuals with the lowest (Q1) M10 and highest (Q4) quartiles for L5 and IV were at significantly higher risk for stroke, but a lower RA saw a significant and graded increase in risk. Thus, 24-h rhythm disturbance assessed by RA provides complementary information for stroke risk. Intriguingly, we found that the risk for stroke was highest in those with an extreme L5 midpoint outside of 2:04 AM and 4:38 AM (suggesting very early or late nocturnal activity) and those with a later M10 midpoint between 2:00 PM and 3:26 PM. Assuming that L5 is in the middle of a 10-h in-bed/sleep period, the finding of L5 timing may be interpreted as increased risk for participants who sleep relatively early (before 9 PM) or later (after 2:30 AM). Similarly, later and extremes of M10 midpoint timing, i.e., being active too early or too late, is associated with increased stroke risk. It is also well-known that stroke sub-types exhibit 24-h rhythmic variation in the time of onset, peaking in the morning hours.^52^ This may relate to risk factors contributing to thrombosis risk similar to observations with myocardial infarctions. ^53^ While we do not have the time of stroke diagnosis in this study, these findings provide evidence for the importance of optimal daily timing for behaviors such as exercise, rest, and sleep according to circadian rhythms^54^ in potential stroke prevention. Future work is needed to examine how rest-activity rhythms interact with the 24-h variation of thrombosis risk.

### Differences between ischemic and hemorrhagic strokes

The association between suppressed 24-h rest-activity rhythms and stroke risk persisted when considering only ischemic stroke but was non-significant in hemorrhagic stroke events. One explanation might be a lack of power due to fewer hemorrhagic stroke events (290), i.e., less than half of that of ischemic stroke events (765). Note that the effects of lower RA and M10 on the risk of hemorrhagic stroke appeared to be in the same direction as that for all strokes or ischemic stroke. A larger sample size is needed to confirm or refute this possibility. Another possibility is that circadian disturbances lead to pathological conditions or risk factors favoring ischemic stroke, including increased cardiovascular risk factors.^55^ This directly impacts stroke risk and recovery and may also indirectly impact daily life cycles that disturb the circadian system leading to a vicious cycle favoring ischemic stroke. For instance, obesity, diabetes, and hypertension are associated with a greater increase in the risk for ischemic stroke than that for hemorrhagic stroke;^56–58^ and circadian disturbances are linked to all these adverse health conditions.^10–12,14^

### Post-stroke outcomes

In the general population, 24-h rhythm disturbances have been linked to many adverse health outcomes, such as the increased risk for disability, neurodegenerative diseases, dementia, and mortality.^21,59,60^ A recent human study also showed that disrupted rest-activity rhythms are associated with a greater cerebral small vessel disease burden in older adults.^40^ The current study supports the role of 24-h rhythm disturbances in post-stroke outcomes, showing a higher risk for dementia, depression, disability, or mortality in those stroke patients with more suppressed 24-h rest-activity rhythms. These results are consistent with the findings in animal studies that circadian disruption worsened stroke outcome (e.g., infarct volume or lesion size) ^61,62^ and increases mortality after ischemic stroke.^63^ The direct pathological pathways through which circadian disturbances impact stroke outcomes remain unclear. Animal studies suggested that circadian disruption impacts stroke outcomes likely through its influence on the inflammatory response, contributing to neurotoxicity and increased neuronal damage in the brain.^61,62^ Further studies are warranted to test this hypothesis in humans.

### Limitations

The strengths of this study include large sample size, objective assessment of 24-h rhythm disturbances, and prospective design with nearly eight years of follow-up. There are also certain noticeable limitations in the study design. One limitation is that we rely on the diagnostic code and may suffer from misclassification and/or underreporting of strokes. Most participants in the UK Biobank are Caucasian of European descent, such that the generalizability of the findings of this current study should be examined in follow-up studies. In addition, UK Biobank participants are healthier than the general UK population, especially those included in this study who volunteered to undergo actigraphy assessment.^37^ Such recruitment bias may lead to underestimation of the observed associations of 24-h rest-activity rhythms with stroke risk and post-stroke outcome because these participants may likely have more ‘normal’ daily activity patterns, fewer comorbidities, lower rates of stroke, and adverse outcomes. This may have biased the associations to the null. Moreover, 6 days for actigraphy assessment may not be long enough to accurately capture the mean and variance of daily activity patterns. The patterns may also change significantly over time during the follow-up. This may have more random influences on the associations of 24-h rest-activity rhythms with stroke outcomes because (1) there was a time lag between actigraphy assessment and incident stroke, and (2) circadian regulation and scheduled daily activities might be different before a stroke and after stroke. Finally, though we controlled for a range of confounders and other risk factors for stroke or adverse health outcomes after stroke, the related variables were assessed at baseline. These factors/variables may change over time (after actigraphy assessment) and serve as the mediators or pathological pathways through which circadian disturbances impact the risk for stroke or stroke outcomes. For example, a non-obese participant with 24-h rhythm disturbances at baseline might become obese, leading to an increased risk of stroke later. Therefore, longitudinal monitoring of 24-h rhythm disturbances and different pathological changes is needed to clarify their temporal interactions and relevance to stroke risk and recovery.

## Conclusion

Suppressed 24-hour amplitude in the daily motor activity rhythm was associated with increased risk of stroke and predicted stroke outcomes, especially mortality, in middle-to-older adults, independent of other known risk factors for stroke and adverse health outcomes. Monitoring of ambulatory daily motor activity with wearable devices may provide a unique opportunity for the assessment of stroke risk. Circadian disturbances may be a modifiable risk factor that can be a target for stroke prevention or stroke rehabilitation.

## Acknowledgments

This research has been conducted using the UK Biobank Resource under Application Number 33883. Dr. L Gao is funded by the National Institutes of Health (NIH) grant R03AG067985 and the Foundation for Anesthesia Education and Research (FAER). Dr. Li is funded by the BrightFocus Foundation Alzheimer’s Disease Research Program A2020886S. Dr. Hu is funded by NIH grants RF1AG059867 and RF1AG064312. Dr. Scheer is funded by the NIH grants R01HL140574 and R01HL153969. The organizations mentioned above had no direct input on this work.

## Author Contributions

Conception and design of the study: Gao, Li, Nogueira, and Hu Acquisition and analysis of data: Gao, Li, Zheng, Nason, and Hu. Drafting a significant portion of the manuscript or figures: Gao, Zheng, Scheer, Nogueira, and Hu.

